# First-in-human prospective trial of sonobiopsy in glioblastoma patients using neuronavigation-guided focused ultrasound

**DOI:** 10.1101/2023.03.17.23287378

**Authors:** Jinyun Yuan, Lu Xu, Chih-Yen Chien, Yaoheng Yang, Yimei Yue, Siaka Fadera, Andrew H. Stark, Katherine E. Schwetye, Arash Nazeri, Rupen Desai, Umeshkumar Athiraman, Aadel A. Chaudhuri, Hong Chen, Eric C. Leuthardt

## Abstract

Sonobiopsy is an emerging technology that combines focused ultrasound (FUS) with microbubbles to enrich circulating brain disease-specific biomarkers for noninvasive molecular diagnosis of brain diseases. Here, we report the first-in-human prospective trial of sonobiopsy in glioblastoma patients to evaluate its feasibility and safety in enriching circulating tumor biomarkers. A nimble FUS device integrated with a clinical neuronavigation system was used to perform sonobiopsy following an established clinical workflow for neuronavigation. Analysis of blood samples collected before and after FUS sonication showed enhanced plasma circulating tumor biomarker levels. Histological analysis of surgically resected tumors confirmed the safety of the procedure. Transcriptome analysis of sonicated and unsonicated tumor tissues found that FUS sonication modulated cell physical structure-related genes but evoked minimal inflammatory response. These feasibility and safety data support the continued investigation of sonobiopsy for noninvasive molecular diagnosis of brain diseases.

## INTRODUCTION

The diagnostic evaluation of glioblastoma (GBM) relies on neuroimaging by magnetic resonance imaging (MRI) and computed tomography, followed by surgical resection or biopsy for histological confirmation and genetic characterization. Alternative approaches to obtain information on a brain lesion without surgery include lumbar puncture and blood draw^1^. Lumbar puncture for cerebral spinal fluid-based liquid biopsy is uncomfortable and carries procedural risk, limiting its use for repeated testing. In contrast, blood-based liquid biopsy is a noninvasive, rapid, and inexpensive method to obtain highly relevant information about the tumor^2^. This approach detects circulating tumor-derived biomarkers, such as DNA, RNA, proteins, and extracellular vesicles shed by tumor cells. It is a promising approach for the diagnosis, molecular characterization, and monitoring of brain tumors^3^. Although blood-based liquid biopsy-guided personalized therapy has already entered clinical practice to treat several cancers^4, 5^, extending it to brain cancer remains challenging^6^. Brain tumor-derived circulating tumor biomarkers are generally detected only at low abundance and in a limited number of patients, which makes analysis difficult in routine clinical practice^7–10^. This low abundance is primarily due to the blood-brain barrier (BBB), a physical barrier that prevents the transfer of brain tumor biomarkers into the peripheral circulation, resulting in low test sensitivity^6, 11^. Even when the BBB is disrupted in GBM, the release of tumor-specific biomarkers into the peripheral circulation remains limited^1^.

Transcranial low-intensity focused ultrasound (FUS) in combination with intravenously injected microbubbles is a promising technique for noninvasive, spatially targeted, and reversible disruption of the BBB^12^. FUS can penetrate the skull noninvasively and focus on virtually any brain region with millimeter-scale accuracy. Microbubbles, traditionally used as blood-pool contrast agents for ultrasound imaging, amplify and localize FUS-mediated mechanical effects on the vasculature via FUS-induced cavitation (i.e., microbubble expansion, contraction, and collapse). Microbubble cavitation generates mechanical forces on the vasculature^13^ and reversibly increases the BBB permeability in the FUS-targeted brain region. Typically, the permeabilized BBB usually reseals after a few hours^14^. Recent clinical studies have demonstrated the feasibility and safety of FUS-mediated BBB disruption for brain drug delivery in patients with glioblastoma^15–20^, Alzheimer’s disease^21^, amyotrophic lateral sclerosis^22^, and Parkinson’s disease^23^

We hypothesized that FUS-induced BBB disruption enables “two-way trafficking” between the brain and bloodstream^24^. With FUS-mediated BBB disruption, circulating agents can enter the brain, while brain tumor-derived biomarkers can be released into the bloodstream for potential diagnostic access. We term this FUS-induced release of biomarkers into the blood circulation for blood-based liquid biopsy as sonobiopsy. Sonobiopsy disrupts the BBB at the spatially targeted brain location, releases tumor-derived biomarkers from precisely defined tumor locations into the blood circulation, and enables timely detection of biomarkers in the blood to minimize clearance. Our previous study provided compelling preclinical evidence that sonobiopsy enriched circulating RNA, DNA, and proteins in small and large animal models^24–28^. Recently, we found that sonobiopsy improved the detection sensitivity of GBM-specific EGFRvIII mutation from 7.14% to 64.71% and TERT C228T from 14.29% to 45.83% in a mouse GBM model. It also improved the diagnostic sensitivity of EGFRvIII from 28.57% to 100% and TERT C228T from 42.86% to 71.43% in a porcine GBM model^29^. By retrospectively analyzing blood samples collected from FUS-mediated drug delivery clinical trials, Meng et al. provided preliminary clinical evidence that FUS-induced BBB disruption increased the concentrations of circulating biomarkers, including cell-free DNA, neuron-derived extracellular vesicles, and brain-specific protein^30^.

The most widely used FUS device in current brain drug delivery clinical trials is the MRI-guided FUS system, ExAblate Neuro, from InSightec Inc. This system utilizes a hemispherical-shaped FUS transducer with 1,024 elements and an aperture of 30 cm^31^. It was initially designed for thermal ablation and has been approved by the United States Food and Drug Administration to treat essential tremors. While this device can be adapted for sonobiopsy, it is expensive (>$3M) and requires MR-compatible hardware, MR scanner time, and extensive training to operate the device. Although neuronavigation-guided FUS devices were developed for drug delivery, they need a robotic arm for positioning heavy FUS transducers with large apertures and customized optical trackers to guide the positioning of the transducer. FUS devices used for drug delivery require high spatial precision and a large treatment volume to deliver drugs to cover the whole diseased brain region efficiently. However, FUS devices for sonobiopsy do not need to deliver therapeutic drugs or cover the entire tumor. Affordable and easy-to-use FUS devices are needed for sonobiopsy to target specific regions inside the tumor for spatially targeted biomarker release.

Here, we present a small-aperture FUS device that is nimble and easily integrated with existing clinical neuronavigation systems. This device enables the sonobiopsy procedure to be performed using a clinical workflow similar to existing neuronavigation-guided tissue biopsy. A pilot prospective sonobiopsy clinical study was conducted on GBM patients using this device to evaluate the feasibility and safety of sonobiopsy. Our results demonstrate that sonobiopsy enriched circulating GBM-specific biomarkers without causing any evident tissue damage.

## RESULTS

### Study patients

The aim of this prospective single-arm trial was to assess the feasibility and safety of sonobiopsy in patients with GBM. The trial was approved by the Washington University in St. Louis Institute Review Board and registered with clinicaltrial.gov (Identifier: NCT05281731). Written informed consent was obtained from all participants prior to study enrollment. Patients with a lesion in the brain with imaging characteristics consistent with GBM were screened for the clinical trial. Of the five patients screened for the study, three patients (two men and one woman; median age 65 years; range 58–74 years) met the inclusion/exclusion criteria and were enrolled in the trial (**Table 1**, **Fig. 1a**). Details of the inclusion and exclusion criteria are provided in Supplementary Table S1. The primary outcome of this study was to evaluate the feasibility of sonobiopsy to increase GBM tumor-specific biomarker levels in the post-sonication blood samples compared with pre-sonication blood samples. The secondary study outcome was to verify that there was no evidence of brain tissue injury associated with the procedure.

**Figure 1.**
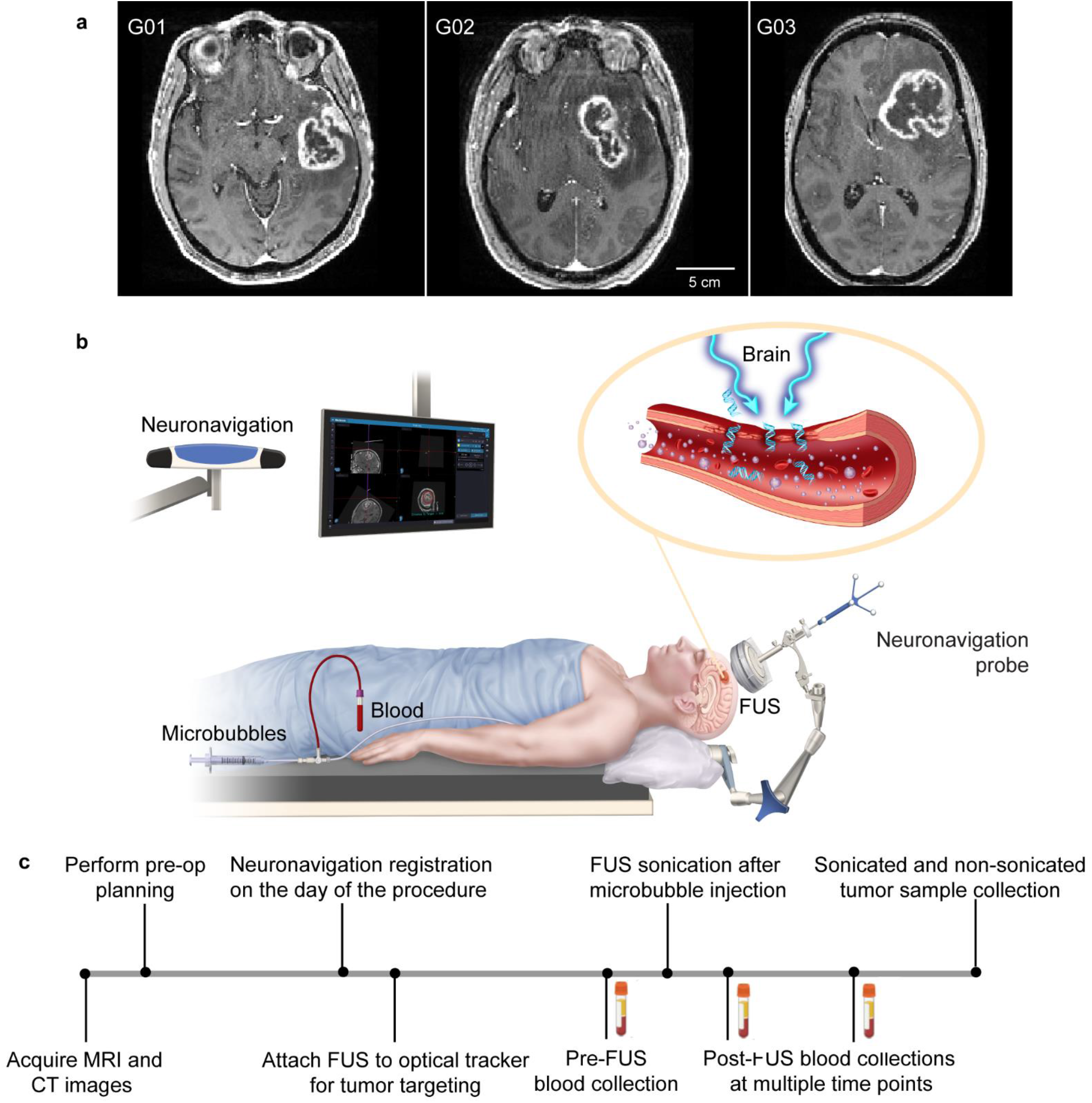
Sonobiopsy procedure. **(a)** Contrast-enhanced T1w MRI images prior to sonobiopsy for three patients (G01, G02, and G03) enrolled in this study. **(b)** Illustration of the neuronavigation-guided sonobiopsy setup. **(c)** Sonobiopsy clinical workflow.

**Table 1.** Patient demographics.

| Patient | Sex | Age range | Diagnosis | IDH-1 | TERT |
| --- | --- | --- | --- | --- | --- |
| G01 | M | 71–75 | Glioblastoma | Wild type | TERT C228T |
| G02 | M | 56–60 | Glioblastoma | Wild type | TERT C250T |
| G03 | F | 66–70 | Glioblastoma | Wild type | TERT C228T |

### Sonobiopsy procedure was successful

Sonobiopsy was performed after the patients were prepared for the surgery in the operating room and before the planned surgical removal of the GBM tumor. Patients were under general anesthesia, and vital signs were continuously monitored by an anesthesiologist. Sonobiopsy was performed using a neuronavigation-guided FUS transducer (**Fig. 1b**). The timeline of the procedure is illustrated in **Fig. 1c**. MRI and CT images acquired before the procedure were loaded in the neuronavigation system (Stealth S8, Medtronic) and used for spatial registration of the patient’s head position. The patient’s hair above the tumor region was shaved. Degassed ultrasound gel was applied to the cleaned scalp for acoustic coupling. The FUS transducer with a water bladder attached was coupled to a standard neuronavigation probe with a customized adaptor (**Fig. 1c**). The focus position of the FUS transducer was calibrated beforehand to be 80 mm from the tip of the stereotactic probe. An 80 mm offset was added in the neuronavigation software so that the tip of the “virtual probe” indicated the location of the FUS focus (**Fig. 2a**). The FUS transducer was mechanically positioned to align its focus at the planned tumor location. The acoustic pressure field was simulated based on the final trajectory of the probe, and the skull attenuation was estimated based on the simulation (**Fig. 2b**). The offset between the planned target and the actual target based on the simulation was found to be 1.91 ± 0.97 mm in lateral direction and 5.29 ± 0.83 mm in the axial direction. The acoustic output pressure of the FUS transducer was adjusted to control the mechanical index (in situ acoustic pressure/square root of frequency) to be within 0.4–0.8 (**Table 2**). Microbubbles (Definity, 10 µL/kg) were intravenously injected by an anesthesiologist, followed by FUS sonication for 3 mins.

**Figure 2.**
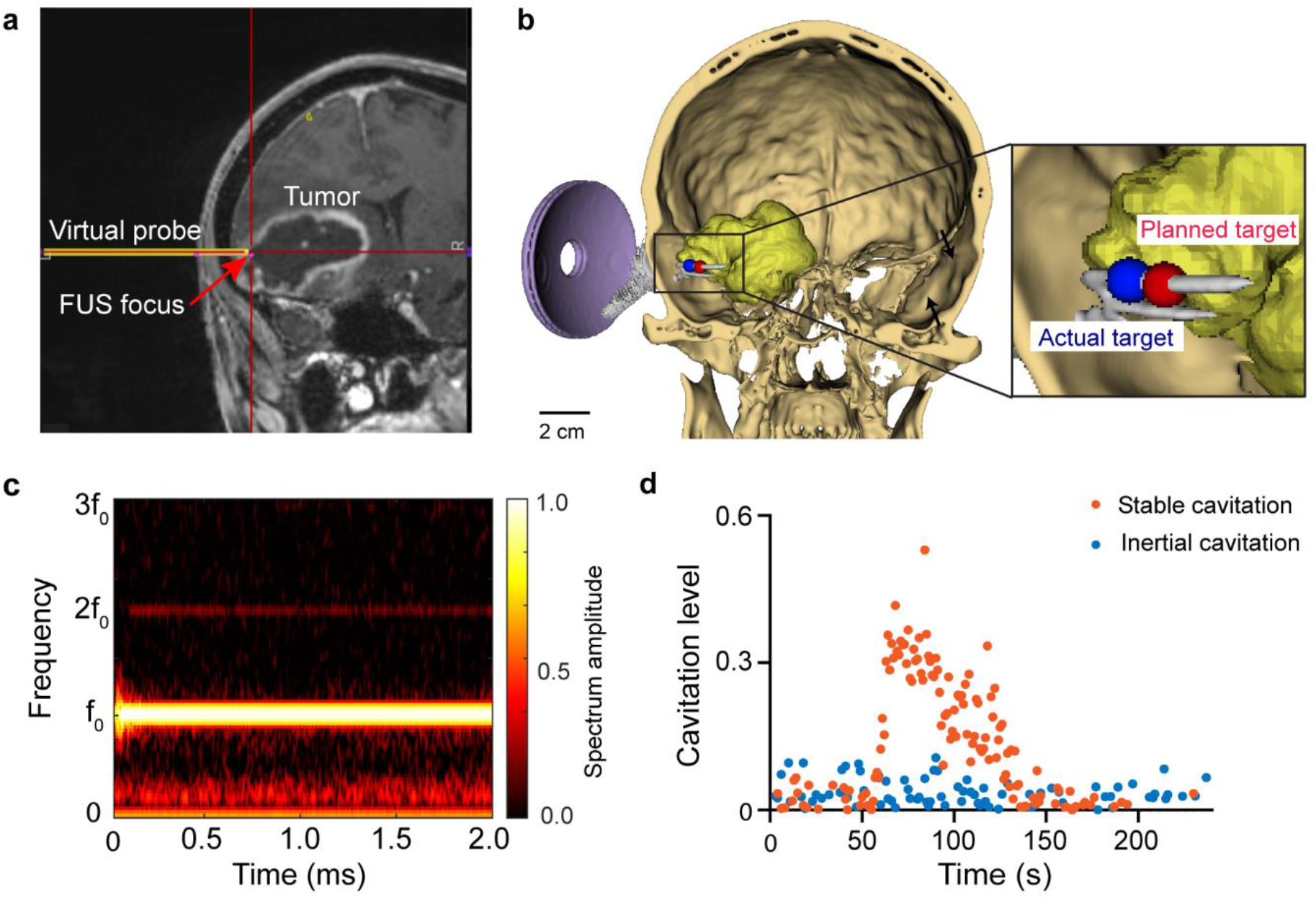
Tumor targeting and treatment monitoring. **(a)** Screenshot from the Stealth neuronavigation system to show the precise alignment of the FUS focus (arrow) with the planned target indicated by the crossing point of the red lines. **(b)** 3D reconstruction to show the spatial location of the planned target inside the tumor and the simulated FUS focus location based on the trajectory obtained from the neuronavigation system. **(c)** Representative time-frequency analysis of the acquired cavitation single during FUS sonication to show the spectrum of the signals. **(d)** Representative stable cavitation and inertial cavitation levels measured based on the frequency spectrum of the acquired cavitation signals.

**Table 2.** Summary of sonobiopsy parameters.

| Patient number | G01 | G02 | G03 |  |
| --- | --- | --- | --- | --- |
| Simulated in situ pressure (MPa) | 0.49 | 0.31 | 0.66 |  |
| Estimated mechanical index (MI) | 0.61 | 0.38 | 0.82 |  |
| Procedure duration from the time when the patient was ready to the end of FUS sonication (min) | 24 | 14 | 30 |  |
| Microbubble dose ($\mu\text{L}/\text{Kg}$) | 10 | 10 | 10 | |
| Microbubble volume (mL) | 5.3 | 8.2 | 4.54 |  |
| Stable cavitation dose (a.u.) | 4.2 | 1.9 | 6.9 |  |
| Inertial cavitation dose (a.u.) | 0.41 | 0.39 | 79.9 |  |

The FUS transducer had an acoustic sensor inserted in its center. The sensor had three functions: quality assurance to ensure the FUS transducer had consistent output before the procedure (**Fig. S1a, S1b**), acoustic coupling quality assessment by performing cavitation detection during FUS sonication before microbubble injection (**Fig. S1c, S1d**), and FUS treatment monitoring after microbubbles were injected (**Fig. 2c, 2d**). Real-time cavitation monitoring provided an effective tool for monitoring the treatment procedure. The stable and inertial cavitation levels were quantified based on the frequency spectrum of the acquired signals to quantify the bubble activity under stable oscillation (stable cavitation generates harmonic signals) and violent collapse (inertial cavitation generates broadband signals). The cavitation doses calculated by integrating the cavitation level over time for all three patients are summarized in **Table 2**.

Blood samples were collected immediately before (5 mins pre-FUS) and at different time points post-sonication. After the last blood collection, surgery was performed, and tumor tissue samples were collected from the FUS sonicated and nonsonicated tumor regions under the neuronavigation guidance. The total procedure time from when the patient was prepared and ready for the sonobiopsy procedure to the end of FUS sonication was 22.7 ± 6.6 min.

### Sonobiopsy enriched circulating GBM-specific biomarkers

To evaluate the feasibility of sonobiopsy in enriching circulating GBM biomarkers, we first quantified plasma levels of glial fibrillary acidic protein (GFAP), a reliable and commonly used liquid biomarker for GBM^32^. Using ultrasensitive single-molecule array (Simoa) assay, we found that sonobiopsy increased plasma GFAP levels for all three patients (**Fig. 3a**), with the maximum increase being 1.2-fold for G02 at 30 mins post-FUS, from 15.6 ±0.83 ng/mL to 19.3 ± 2.4 ng/mL of plasma (p<0.05). We further analyzed the total cell-free DNA (cfDNA) levels in the plasma (**Fig. S2)**. We found that sonobiopsy significantly increased the concentration of mono-nucleosomal cfDNA fragment (120–280 bp) for all time points post-FUS, except at 5 mins for G01 (**Fig. 3b**). The maximum increase was 1.6-fold for G02 at 30 mins, from 30.3 ± 4.2 ng/mL to 63.9 ± 4.6 ng/mL of plasma (p<0.0001).

**Figure 3.**
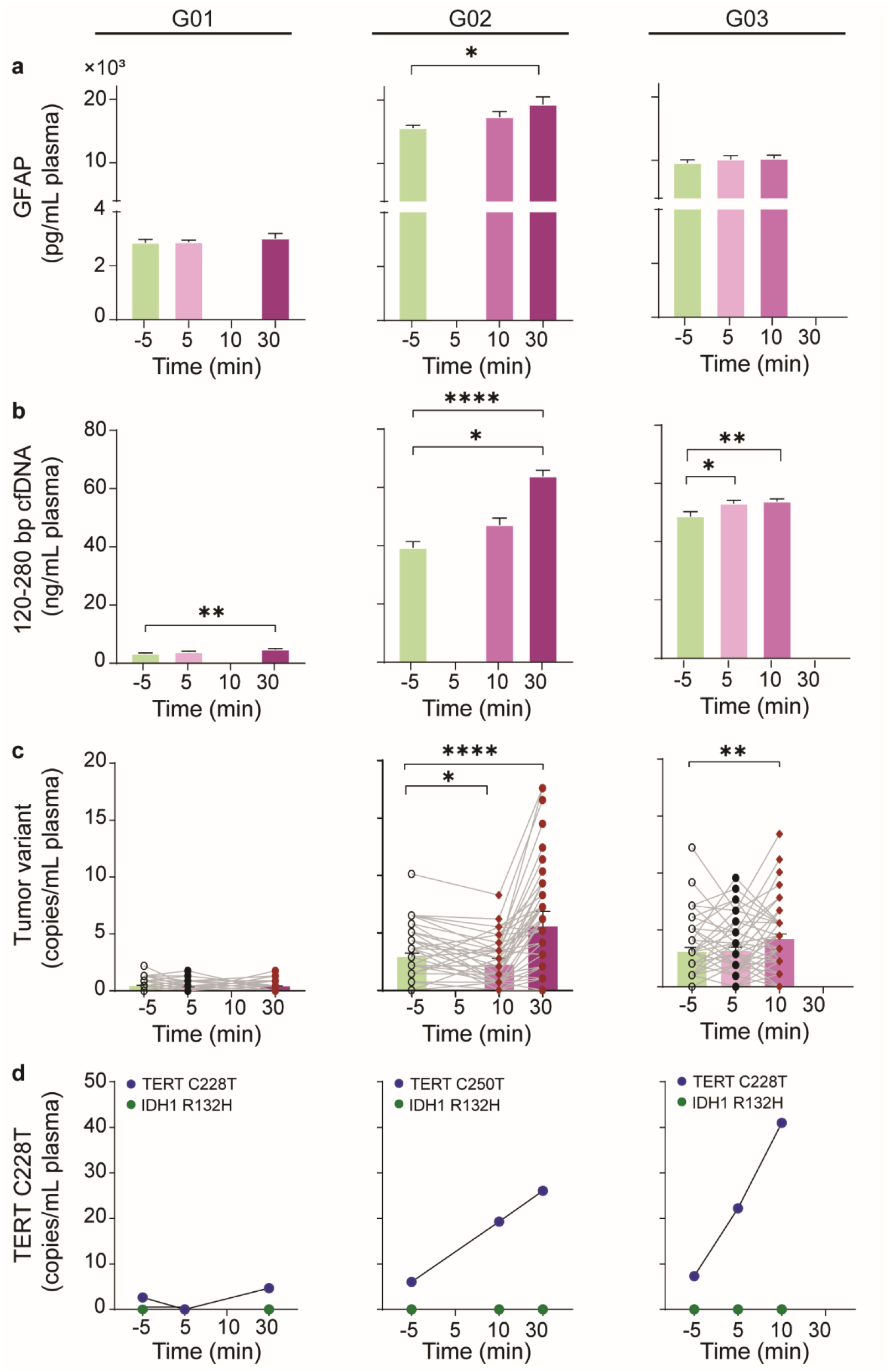
Sonobiopsy enriched circulating GBM-specific biomarkers. **(a)** Plasma GFAP concentration measured at 5 mins pre-FUS (-5 min) and different time points post-FUS (5, 10, and/or 30 mins) for three patients (G01, G02, and G03) [unpaired parametric *t*-test comparing post-FUS each time point with pre-FUS, * p<0.05. ** p<0.01. *** p<0.001. ***** p<0.0001, bar graph represents mean ± stand deviation (SD)]. **(b)** Concentration of single nucleosome length cfDNA (120–280 bp fragments) was significantly increased after FUS for all three patients (unpaired parametric *t*-test comparing post-FUS time points with pre-FUS). **(c)** Patient-specific tumor variants detected using a personalized tumor-informed ctDNA assay were significantly increased for G02 and G03 (paired parametric *t*-test comparing post-FUS time points with pre- FUS). **(d)** ddPCR analysis of plasma TERT and IDH1 shows sonobiopsy increased the level of TERT mutation, but not the wild type IDH1.

To assess the potential of sonobiopsy to improve the detection of patient-specific tumor variants in the plasma, we utilized a personalized tumor-informed ctDNA assay (Invitae Personalized Cancer Monitoring assay). This assay involved performing whole exome sequencing (WES) on the tumor and normal tissue. Sequencing results allowed the identification and selection of up to 50 clonal, somatic, single-nucleotide variants present in the tumors but not the matched normal samples. The selected single-nucleotide variants were used in the design of a patient-specific panel, which was used to detect circulating tumor DNA (ctDNA) in patient plasma samples. We compared the absolute level of patient-specific tumor variants in the plasma samples collected before and after FUS using tumor variant copies/ml plasma. Our results indicated that sonobiopsy successfully enhanced the detection of patient-specific tumor variants in the plasma of G02 and G03 (**Fig. 3c**). For these two patients, the kinetics of biomarker changes following sonication revealed that higher tumor variant copies were detected at later time points. Specifically, G02 reached the highest number of tumor variant copies at 30 min post-FUS, compared with pre-FUS (5.6 vs. 2.9 for post vs. pre-FUS, p<0.001). For G03, the final blood sample was acquired at 10 mins post-FUS, and the tumor variant copies were the highest at this time point compared with pre-FUS (4.2 vs. 3.1 for post vs. pre-FUS, p<0.01). However, no clear increase in tumor variant copies was observed for G01.

A combination of TERT promoter mutation and IDH wild type is the most common genotype observed in GBM. TERT promoter mutations are present in 62% of GBM patients and associated with poor treatment outcome^33, 34^, while IDH1 mutations are also important diagnostic and prognostic markers for glioma^35^.To further investigate the potential of sonobiopsy in discerning these two mutations, we analyzed the amount of TERT mutation (C228T and C250T) and IDH1 mutation (R132H) in the plasmas with digital droplet PCR (ddPCR). ddPCR analysis on tumor tissue samples found that all three patients were positive for TERT mutations but IDH1 wild type (**Fig. S3**). IDH1 mutation (R132H) was consistently undetectable in any of the sonobiopsy plasma samples. In contrast, TERT mutations (C228T and C2250T) were detected at higher levels by sonobiopsy than with conventional blood draw pre-FUS, with 1.8 fold increase for G01 at 30 mins post-FUS, 4.3-fold increase for G02 at 30 mins post-FUS, and 3.0-fold increase for G03 at 10 mins post-FUS (**Fig. 3d**).

### Sonobiopsy did not induce detectable tissue damage

During the FUS sonication procedure, we did not observe any significant fluctuations in vital signs, such as heart rate and respiration, nor did we note any adverse events. Following FUS sonication, we observed no evidence of hemorrhage on the surface of the brain due to FUS sonication (**Fig. 4a**). Additionally, tissue samples obtained from both the sonicated and nonsonicated tumor regions after tumor dissection were stained by hematoxylin and eosin (**Fig. 4b**). Signs of tissue damage, including microhemorrhages and other cytoarchitectural changes, were not detected.

**Figure 4.**
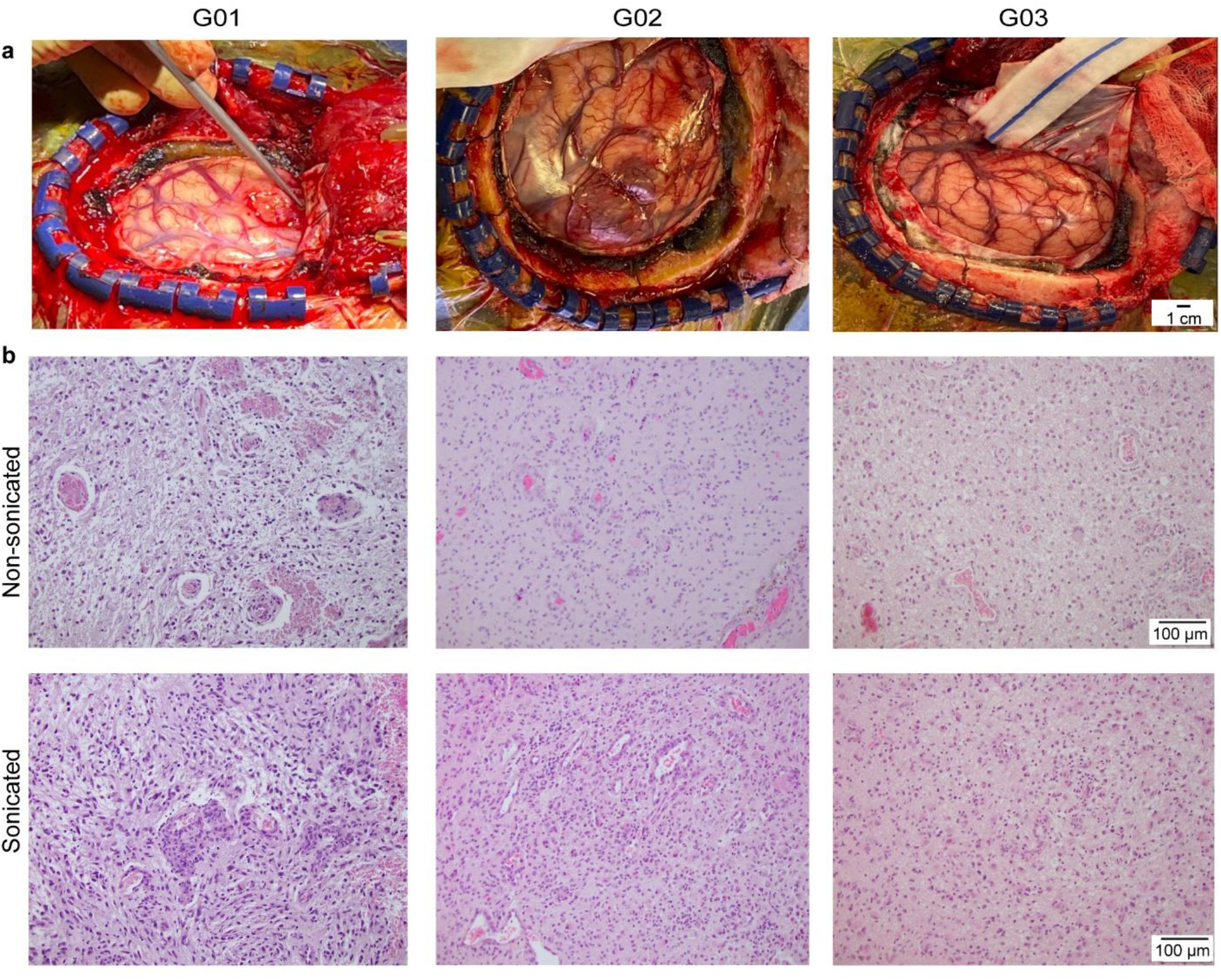
Safety of sonobiopsy by tissue analysis. **(a)** Gross pathology examination of the surface of the brain after craniotomy for the three patients (G01, G02, and G03) did not observe tissue damage induced by the FUS procedure. **(b)** Hematoxylin and eosin (H&E) staining of the sonication and nonsonicated brain tumor tissue did not observe clear evidence of tissue damage.

### Sonobiopsy did not induce evident inflammation/immune responses

To provide a comprehensive safety profile of FUS sonication on the tumor, we conducted transcriptome analysis of sonicated and nonsonicated tumor tissues. We identified differentially expressed genes (DEGs) using hierarchical clustering analysis following strict criteria of the absolute value of log2 (fold-change) > 2 and Pvalue < 0.05 (**Fig. 5a**). Our analysis identified 34 DEGs out of the total 17,982 genes (0.19%), among which 19 transcripts were identified as upregulated DEGs associated with sonication, while 15 were downregulated DEGs (**Fig. 5b**).

**Figure 5.**
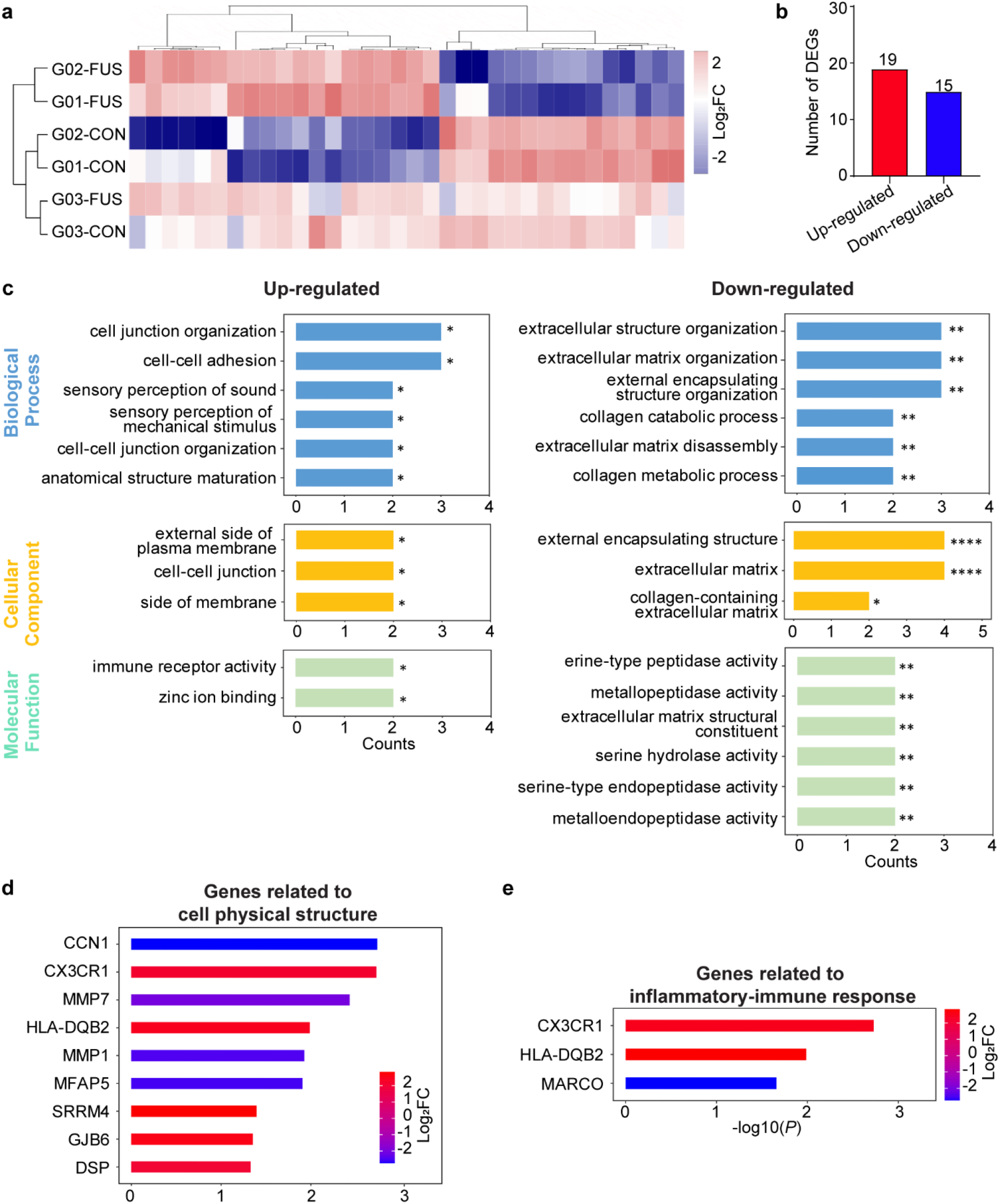
Transcriptome analysis of differentially expressed genes (DEGs) after sonobiopsy. **(a)** Hierarchical clustering heat map of all DEGs with log2FC (fold change) values > 2 and p< 0.05. Rows represent brain tissue samples acquired from the FUS-sonicated (FUS) or nonsonicated (CON) tumor regions from different patients, and columns represent individual DEGs. **(b)** Summary of the total number of upregulated and downregulated DEGs. **(c)** Upregulated (red) and downregulated (blue) DEGs with GO functional analysis for enriched biological processes, cellular components, and molecular functions. *p<0.05, **p<0.01, ***p<0.001, ****p<0.0001. **(d)** Upregulated and downregulated genes involved in GO terms related to cell physical structure. **(e)** Upregulated and downregulated genes involved in GO terms related to inflammatory or immune-related response.

The gene ontology (GO) analysis of the upregulated and downregulated DEGs showed that the enriched GO terms were related to the physical structures of cells, including their interactions with neighboring cells and their surrounding extracellular matrix (**Fig. 5c**). This suggests that FUS- combined with microbubbles caused mechanical perturbation to the cell-cell and cell-matrix interaction. The genes related to cell physical structure that were upregulated and downregulated are further summarized in **Fig. 5d**. Real-time qRT-PCR analysis was performed and verified the upregulation and downregulation of these genes (**Fig. S4**).

FUS with microbubbles was reported by previous studies to induce sterile inflammation in healthy mouse brains^36–38^. However, our analysis only identified one immune-related GO term, “immune receptor activity,” among the top 26 enriched GO terms (**Fig. 5e**). Further analysis of potential inflammatory-immune-related GO terms involving at least one DEG identified only one significant GO term (GO0140375, “immune receptor activity”) that had more than one overlapped DEGs. Among all the discovered GO terms, CX3CR1 and HLA-DQB2 were identified as upregulated DEGs, and MARCO was identified as a downregulated DEG. The upregulation of CX3CR1 and downregulation of MARCO were further confirmed by real-time qRT-PCR results (**Fig. S4**). Our results suggest that the FUS procedure did not induce severe immune or inflammation responses.

## DISCUSSION

We present the first prospective clinical study of sonobiopsy in GBM patients. Our study utilized the nimble sonobiopsy device, which was seamlessly integrated with an existing clinical neuronavigation system. The sonobiopsy procedure was performed using an established clinical workflow for neuronavigation. Our findings provide crucial initial evidence that sonobiopsy can enrich GBM biomarkers in the blood by targeting specific tumor locations and coordinating the blood collection time.

We demonstrated significant technological advancements that the sonobiopsy device offers to adopt this innovative technique in the clinic. First, the nimble design of the FUS device allows for direct attachment of the FUS device to existing neuronavigation probe used by any clinical neuronavigation system, enabling precise positioning of the FUS transducer with high accuracy. Furthermore, this unique design also allows for easy integration of sonobiopsy into the existing clinical workflow, eliminating the need for additional training of neurosurgeons to perform the sonobiopsy procedure. This will reduce the barrier to adopting this technique in the future. Importantly for future considerations, an operating room is not required. While cranial fixation and anesthesia were used for the patients described in this work, it is not essential. Standard navigation techniques that can be used without fixation and anesthesia could enable sonobiopsies to be performed outside classic operative and procedural environments (e.g., hospital rooms and clinics). Second, the sonobiopsy device also incorporates numerical simulation of the acoustic energy delivered into the brain. This simulation provides critical guidance for the selection of ultrasound parameters and allows visualization of the ultrasound beam shape and location inside the brain. Third, the integration of the FUS device with cavitation detection enables the development of quality assurance for the FUS device and real-time monitoring for the treatment. Future work can integrate cavitation feedback control algorithms to regulate the FUS acoustic pressure in real-time^39, 40^, which would ensure precise and consistent delivery of the ultrasound energy.

We also demonstrated that sonobiopsy could be integrated with advanced blood-based biomarker analysis techniques for the noninvasive and spatially targeted molecular diagnosis of GBM without causing brain damage. ctDNA-based sequencing assays can be divided into two classes: tumor-naïve assays and tumor-informed assays. Tumor-naïve assays use broad panel-based sequencing assays for genotyping or tumor early detection with a detection limit of about 0.2%^41^; Tumor-informed assays are designed in reference to mutations known from the tumor and can reach a limit of detection as low as 0.01% variant allele frequency^41^. Examples of tumor-informed assays include CAPP-Seq^42^, PhasED-seq^43^, and personalized tumor-specific sequencing^43^. In this study, we used the Invitae Personalized Cancer Monitoring assay, which was developed to detect residual molecular diseases by providing sensitive detection of ctDNA in the plasma samples. Our results show that sonobiopsy significantly increased the detection of tumor variants in two out of three patients. Given that this is the first prospective trial, the observation that 2/3 of patients had a significant increase in the detected tumor variants suggests that sonobiopsy is a promising technique for enriching plasma ctDAN. In addition to sequencing-based assays, ddPCR is a targeted approach for rapidly detecting specific known mutations with high sensitivity and tissue concordance^44–46^. Thus, ddPCR was used in our study to detect ctDNA with prior knowledge of the mutations expressed by the GBM tumors. The GBM tumors are known to have TERT mutation but no IDH1 mutation. The ddPCR results demonstrate that sonobiopsy enriched the level of TERT mutation without affecting the amount of IDH1 mutation, implying that sonobiopsy can improve the sensitivity and specificity in mutation detection. The kinetics of biomarker changes post-FUS indicate a trend of increasing ctDNA levels within the 5–30 min time frame. Future studies are needed to determine the complete kinetics of biomarker release and optimal blood collection time.

The prospective trial design implemented in this study allowed for the collection of GBM tissue samples from the sonicated and nonsonicated tumor regions in each patient. This approach provided an unprecedented opportunity to evaluate the bioeffects of FUS sonication on the tumor in GBM patients. We conducted the transcriptome analysis of FUS effects on patient GBM tumors obtained at 70.0 ± 6.1 minutes post-FUS sonication and observed only a minimal change of 0.19% in gene expression after sonication. Most upregulated and downregulated genes were related to the physical structures of cells, such as cell interactions with neighboring cells and the extracellular matrix. This finding suggests that FUS-combined with microbubbles caused mechanical perturbation to cell-cell and cell-matrix interactions. Notably, the downregulation of MMP1 and MMP7 genes was previously reported to be associated with BBB integrity^47^, suggesting that the sonobiopsy procedure induced BBB disruption at the targeted tumor region. FUS-induced BBB disruption has been shown to induce an inflammatory response in mice^36–38^, but there have been no reports examining the immune response in GBM patients. Contrary to these reports in mice, our study shows that only three genes CX3CR1, HLA-DQB2, and MARCO were identified as DEGs related to the immune response in GBM tumors obtained at 70.0 ± 6.1 minutes post-FUS sonication. CX3CR1 and HLA-DQB2 were upregulated upon FUS sonication, consistent with previously reported increased infiltration of CX3CR1-positive immune cells into the tumor area after FUS-induced BBB disruption in mice^48^. This lack of activation of the immune response needs to be confirmed in the future using tissue samples acquired at later time points.

The results presented in this pilot clinical trial provide important insights into the potential for sonobiopsy in noninvasive molecular characterization of GBM and other brain diseases (e.g., neurodegenerative diseases and psychiatric disorders). Sonobiopsy has the potential to achieve several critical benefits after integration into clinical practice as a complement to neuroimaging and tissue biopsy, including the identification of genetic features before surgical intervention, enabling alterations in surgical strategy. It could also enable the rapid determination of the molecular identity of suspicious lesions observed on neuroimaging scans, particularly in patients who are poor surgical candidates. Furthermore, the ability to repeatedly sample and monitor tumor recurrence and treatment response could provide valuable information to clinicians. In challenging situations where assessment based on neuroimaging alone remains difficult, such as distinguishing treatment-induced pseudoprogression from true relapse, sonobiopsy could provide complementary information. Moreover, it has the potential to support investigations into tumor-specific molecular mechanisms driving disease and accelerate the development of new treatment strategies.

While this study presents milestone achievements in developing sonobiopsy for the molecular diagnosis of GBM, several limitations exist. First, although the data were extremely promising to show the feasibility and safety of sonobiopsy, this pilot study was performed with a small number of GBM patients. Further studies with larger sample sizes are needed to confirm these initial findings and establish the clinical utility of sonobiopsy. Second, we selected to target a single brain location in this pilot study. The previous retrospective study by Meng et al. found that increasing the sonication volume could increase the biomarker release efficiency^30^. Future study is needed to optimize the sonobiopsy procedure, including evaluating the impact of sonication volume on the efficiency of sonobiopsy. Third, there is always a spatial shift in the brain during the surgical dissection of the sonication brain tumor tissue. This potential shift could have introduced an error in the localization of the FUS-sonicated tumor region. To reduce the potential impact of this error, the targeted tumor region was selected to be located at the relative superficial tumor location so that this region was encountered early in the surgery and before substantial brain shift.

In conclusion, this study represents the seminal initial step in establishing the feasibility and safety of sonobiopsy in the brain of patients with GBM. This technique enables noninvasive, spatially targeted, temporally controlled detection of brain GBM biomarkers in the blood. The feasibility and safety data obtained from this pilot study will enable the field to move forward in translating sonobiopsy into impactful diagnostics for GBM and other brain diseases.

## METHODS

### Study design

This prospective, single-arm, single-center, first-in-human study was designed to evaluate the feasibility and safety of sonobiopsy. This study was approved by the Research Ethics Board at Washington University in St. Louis, School of Medicine, and was registered with ClinicalTrials.gov (NCT0528173). All subjects provided written informed consent before enrollment. This trial complied with the International Conference on Harmonization guideline for Good Clinical Practice, Tri-Council Policy Statement on ethical conduct for research involving humans (TCPS-2).

### Sonobiopsy device

A FUS transducer consisting of 15 concentric individual ring transducers with a center frequency of 650 kHz (Imasonics, Voray-sur-l’Ognon, France) was used. The aperture of the transducer was 65 mm, and the focal distance was 65 mm (f-number = 1). The FUS transducer was integrated with a passive acoustic detector at its center. The FUS transducer was driven by a commercial FUS system (Image Guided Therapy, Pessac, France). The nimble FUS transducer was coupled to the passive blunt probe (Stealth S8, Medtronic) through a customized adaptor. The adaptor was attached to the back of the FUS transducer and connected to a cylinder that was aligned with the central axis of the FUS transducer. The diameter of the cylinder matched that of the neuronavigation probe. This adaptor design leveraged the light weight of the FUS transducer and mechanically co-aligned the neuronavigation tracker to the central axis of the transducer. The location of the FUS focus was calibrated to be 80 mm from the tip of the passive blunt probe along the trajectory of the probe. An 80 mm offset was added in the neuronavigation software so that the tip of the “virtual probe” indicated the location of the FUS focus.

### Sonobiopsy clinical workflow

The overall workflow of this clinical study is summarized in **Fig. 1d**. It consists of four main steps: treatment planning, patient preparation, FUS sonication, and blood and tissue collection.

Step 1: Treatment planning. CT and MRI images of the patient’s head were acquired a few days before the procedure. FUS sonication trajectory was planned using the Medtronic S8 planning station (Medtronic Plc, Dublin, Ireland). The trajectory was selected using the following criteria: close to 90° incident angle (best effort), focus depth below skin < 35 mm (limited by the focal length of our FUS transducer), and avoiding ultrasound beam passing through the ear lobe and eye. A full-wave acoustic simulation using the k-Wave toolbox was performed to estimate the ultrasound pressure field distribution inside the brain and calculate the skull attenuation using methods reported in our previous publication^49^.

Step 2: Patient preparation. On the day of the procedure, Mayfield skull clamp (Integra LifeSciences, Princeton, NJ, USA) was fixed to the patient’s head under local and general anesthetic. The skull clamp was then connected to the surgical table through a Mayfield bed attachment. The stealth arc and Veltek arm were then connected, and the patient’s head was registered to the pre-acquired MRI/CT images. The planned FUS trajectory was then entered into the neuronavigation system. A small patch of hair above the tumor region was shaved, and the exposed skin was thoroughly cleaned with alcohol pads. Deionized water was filled into the transducer water bladder, which was continuously degassed with a degassing system for more than 15 min. Degassed ultrasound gel was applied liberally to the exposed skin area. FUS transducer was then placed on the patient’s head under the guidance of the neuronavigation system.

Step 3: FUS sonication. The passive cavitation detector was used to check the quality of the acoustic coupling between the FUS transducer and the skin. If broadband emissions were present in the detect signals when the FUS was turned on without microbubble injection (**Fig. S1**), the most likely cause was due to air bubbles trapped in the coupling media. In this case, we would remove the FUS transducer, clean it, and re-apply the ultrasound gel. The input electrical power was determined based on hydrophone calibration of the FUS transducer focal pressure over different input electrical powers derated by the skull attenuation estimated in Step 1 based on k-wave simulation. To ensure the safety of this study, the estimated in situ acoustic pressure was selected to ensure the mechanical index (MI) was below 0.8, consistent with other FUS-BBBD drug delivery clinical studies^50, 51^. Acoustic parameters besides input power were selected to be the same as our previous preclinical work^29^. The FUS parameters were: center frequency = 0.65 MHz (f_0_); pulse repetition frequency = 1 Hz; pulse duration = 10 ms; treatment duration = 3 min. Fifteen seconds after FUS sonication began, microbubbles (Definity, Lantheus Medical Imaging, North Billerica, MA) were administered intravenously by the standing anesthesiologist at a dose of 10 µL/kg body weight diluted with saline and followed with a saline flush. The injection rate was controlled with the best effort to be 10 s/mL, recommended by the manufacturer. In reference to our previous publication^52^, a custom MATLAB script was written to process the acquired cavitation data to evaluate the stable cavitation and inertial cavitation levels. Briefly, the stable and inertial cavitation levels were calculated as the root-mean-squared amplitudes of subharmonic (f_0_/2 ± 0.15 MHz) and broadband (0.3–2 MHz after removing f_0_/2 ± 0.15 MHz and nf0 ± 0.15 MHz where n = 1, 2, 3) signals, respectively.

Step 4: Blood and tissue collection. Blood samples (20 mL each) were collected 5 mins before and within 30 mins after FUS sonication. Blood samples were stored in BD Vacutainer® EDTA (BD Biosciences, San Jose, California) tubes or Cell-Free DNA BCT (Streck Laboratories, La Vista, Nebraska) tubes. Within 4 hrs of collection, whole blood samples were centrifuged at 1200xg for 10 mins at 4°C. Isolated plasma was centrifuged a second time at 1800xg for 5 mins at 4°C to further remove cell debris. Plasma aliquots were put on dry ice immediately for snap freezing and stored at -80°C subsequently for later downstream analysis. The plasma-depleted whole blood cells were stored as well for tissue sequencing analysis. After blood collection, craniotomy was perforemed and the tumor was resected under the guidance of the neuronavigation system. Sonicated and nonsonicated part of tumor tissue was collected from the resected tumor. Skin tissue on the trajectory of FUS sonication was also collected during surgery.

The collected tissues were fixed in formalin for paraffin embedding or put in fresh medium for snap freezing.

### Plasma protein detection

Frozen plasma samples were thawed at room temperature. All plasma GFAP protein measurements were performed in 2–4 replicates using Simoa^®^ Neurology 2-Plex B Kit on a fully automated HD-X Analyzer (Quanterix, Lexington, MA, United States).

### Cell-free DNA extraction and quantification

Plasma/Serum cfc-DNA/cfc-RNA Advanced Fractionation Kit (Norgen Biotek, Thorold, ON, Canada) was used to extract cfDNA from patient plasma per the manufacturer’s protocol. cfDNA was eluted in 50 µL of each corresponding buffer and was quantified using Qubit Fluorometric Quantitation (Thermo Fisher Scientific). The 2100 Bioanalyzer (Agilent Technologies) was used to assess the size distribution and concentration of cfDNA extracted from plasma samples. The cfDNA in the mononucleosomal size range (120–180 bp) was determined with the software as the area under the peaks^53^.

### Personalized tumor-informed ctDNA assay

Invitae Personalized Cancer Monitoring (PCM) was adopted to detect the patient-specific tumor variants in patients’ plasmas in the following three main steps. First, the data from whole exome sequencing (WES) on tumor and normal (peripheral blood, PB) samples were processed using Invitae’s WES Pipeline. The variants identified from the tumor and normal samples were then compared to identify patient-specific tumor variants. Variant calls were used as input for the minimal residual disease (MRD) Panel Designer pipeline. Second, patient-specific panels (PSPs) were designed to target up to 50 patient-specific single tumor variants. The Panel Designer identified high-confidence patient-specific tumor variants which could be targeted using an Anchored Multiplex PCR (AMP) panel. Third, cfDNAs were extracted from the patient’s plasma samples and used as input for an AMP library preparation using the personalized panel designed for the patient. Libraries were sequenced using the NovaSeq 6000 sequencing platform (Illumina, San Diego, CA, USA) and the resulting fastq files were analyzed using the Invitae MRD calling pipeline. The MRD analysis pipeline aligns MRD library sequences to the genome, calculates the error rates for the targeted variants and measures the allele fraction for the targeted variants. The observed allele fractions are compared to the background error rate to determine the MRD call. To calculate the concentrations of patient-specific tumor variants in each plasma, the value of alternative observations (AOs) from the Inviate MRD was normalized to the input volume of plasma (tumor variant copies/ml plasma).

### ddPCR assays

Custom sequence-specific primers and fluorescent probes were designed and synthesized for patient-specific variant detection (Sigma Aldrich). The forward and reverse primer and probe sequences are listed in **Table S2**. ddPCR reactions were prepared with 2× ddPCR Supermix for probes (no dUTP) (Bio-Rad, Hercules, CA, USA), 2 µL of target cfDNA product, 0.1 µM forward and reverse primers, and 0.1 µM probes. Alternatively, 100 µM 7-deaza-dGTP (New England Biolabs, Beverly, MA, USA) was added to improve PCR amplification for GC rich regions. The QX200 manual droplet generator (Bio-Rad, Hercules, CA, USA) was used to generate droplets. The PCR step was performed on a C1000 Touch Thermal Cycler (Bio-Rad, Hercules, CA, USA) by use of the following program: 1 cycle at 95°C for 10 mins, 48 cycles at 95°C for 30 s and 60°C for 1 min, 1 cycle at 98°C for 10 mins, and 1 cycle at 4°C infinite, all at a ramp rate of 2°C/s. All plasma samples were analyzed in technical duplicate or triplicate based on sample availability. Data were acquired on the QX200 droplet reader (Bio-Rad, Hercules, CA, USA) and analyzed using QuantaSoft Analysis Pro (Bio-Rad, Hercules, CA, USA). All results were manually reviewed for false positives and background noise droplets based on negative and positive control samples. Tumor variant ctDNA concentrations (copies/ml plasma) were calculated by multiplying the concentration (provided by QuantaSoft) by elution volume, divided by the input plasma volume used during cfDNA extraction.

### Bulk RNA sequencing and analysis

Bulk RNA sequencing and analysis were conducted at Genome Technology Access Center at the McDonnell Genome Institute (GTAC@MGI) at Washington University in St. Louis. According to the manufacturer’s instructions, snap-frozen sonicated and nonsonicated tumor tissues were homogenized and isolated using the RNeasy MiniPlus Kit (Qiagen, Hilden, Germany). Total RNA integrity was determined using Agilent Bioanalyzer (Agilent Technologies, Palo Alto, CA, USA). Ribosomal RNA was removed by a hybridization method using Ribo-ZERO kits (Illumina-EpiCentre, San Diego, CA, USA). mRNA was reverse transcribed to yield cDNA using SuperScript III RT enzyme (Thermo Fisher Scientific, Carlsbad, CA, USA) and random hexamers. A second strand reaction was performed to yield double-stranded cDNA. cDNA was blunt-ended, had an A base added to the 3’ ends, and then had Illumina sequencing adapters ligated to the ends. Ligated fragments were then amplified for 12-15 cycles using primers incorporating unique dual index tags. Fragments were sequenced on an Illumina NovaSeq-6000 (Illumina, San Diego, CA, USA) using paired-end reads extending 150 bases. Basecalls and demultiplexing were performed with Illumina’s bcl2fastq2 software. RNA-seq reads were then aligned and quantitated to the Ensembl release 101 primary assembly with an Illumina DRAGEN Bio-IT on-premise server running version 3.9.3-8 software.

All gene counts were then imported into the R/Bioconductor package EdgeR and TMM normalization size factors were calculated to adjust for samples for differences in library size. Ribosomal genes and genes not expressed in the smallest group size minus one sample greater than one count per million were excluded from further analysis. The TMM size factors and the matrix of counts were then imported into the R/Bioconductor package Limma. Weighted likelihoods based on the observed mean-variance relationship of every gene and sample were then calculated for all samples and the count matrix was transformed to moderated log 2 counts-per-million with Limma’s voomWithQualityWeights. The performance of all genes was assessed with plots of the residual standard deviation of every gene to their average log count with a robustly fitted trend line of the residuals. Differential expression analysis was then performed to analyze for differences between conditions and the results were filtered for only those genes with Benjamini-Hochberg false-discovery rate adjusted P values less than or equal to 0.05.

To identify differentially expressed genes (DEGs), we applied strict criteria: log2 (fold-change) > 2 and p< 0.05 for upregulated genes, and log2 (fold-change) < -2 and p< 0.05 for downregulated genes. Gene ontology (GO) enrichment analysis was performed using the g:Profiler tool (https://biit.cs.ut.ee/gprofiler/) on the DEG list classified into upregulated and downregulated lists. The results were visualized as bar plots or dot plots, which were generated in R using ggplot. Raw data were evaluated for statistical significance with a threshold of p<0.05, using an independent *t*-test to compare fold changes.

### Histological analysis

Brain tumor tissue from sonicated and nonsonicated regions were resected and fixed in formalin for paraffin embedding. The brain tumor tissue samples were sectioned to 10 μm slices for hematoxylin and eosin (H&E) staining to examine red blood cell extravasation and cellular injury. Digital images of tissue sections were obtained using an all-in-one microscope (BZ-X810, Keyence, Osaka, Japan). Histological images were assessed by an experienced clinical neuropathologist.

### Statistical analysis

Statistics analysis was performed in Graphpad (Prism) (Graphpad, Boston, MA, USA). Unpaired parametric *t*-test were used to compare the level of GFAP and cfDNA in post-FUS plasmas with that in pre-FUS plasma. Paired parametric *t*-test were conducted to compare the concentrations of tumor variants in post-FUS plasmas with that in pre-FUS plasma. All reported P values are two- tailed unless otherwise specified.

## Data Availability

All data that support the findings of this study are available from the corresponding authors upon reasonable request.

## Acknowledgments

This work was supported by the National Institutes of Health R01CA276174, R01MH116981, UG3MH126861, R01EB027223, and R01EB030102.

## Author contributions

Conceptualization: ECL, HC Experiment design: ECL, HC Experiment implementation: ECL, HC, UA, XL, CYC, JY, SF, AS. Result Investigation: HC, XL, CYC, JY, KS. Funding acquisition: ECL, HC. Project administration: ECL, HC. Supervision: ECL, HC. Writing – original draft: HC, JY, XL, CYC Writing – review & editing: ECL, HC, JY, XL, CYC, YY, SF, AC, KS, AN, RD, UA.

## Competing interests

ECL and HC have a patent on the sonobiopsy technique (US20190323086A1).

## Notes

### Clinical Trial

NCT0528173

### Author Declarations

This study was approved by the Research Ethics Board at Washington University in St. Louis, School of Medicine, and was registered with ClinicalTrials.gov (NCT0528173).

