## Supplementary information for "First-in-human prospective trial of sonobiopsy in glioblastoma patients using neuronavigation-guided focused ultrasound"

**The PDF file includes:**

**Tables S1 and S2**

**Figs. S1 to S4**

**Table S1. Inclusion and exclusion criteria.**

Inclusion Criteria:

- Must be newly diagnosed with a lesion in the brain with imaging characteristics consistent with glioblastoma multiforme. Scan must have occurred no more than 28 days prior to enrollment.
- Lesion must be > 3 cm in maximal dimension on MRI.
- Lesion must be in the supratentorial space within 5 cm of the cortical surface.
- Lesion must be gadolinium enhancing.
- Low grade tumors and metastatic tumors
- Recurrent brain tumors and/or radiation necrosis
- Must be planning to undergo surgical resection of the tumor.
- Must be at least 18 years old.

Exclusion Criteria:

- Contraindication to MRI.
- Previous cranial surgery.
- Previous history of cancer and/or cancer treatments.
- Coagulopathy within 14 days of enrollment defined as PT/PTT outside of normal parameters and platelets < 100,000/mcL.
- Physical skull defect of any kind.
- Ferrous material in the scalp or skull.
- Scalp or skin disease that limits contact with the ultrasound probe.
- Enrolled in another clinical trial where intervention is administered prior to surgery.
- Known hypersensitivity to polyethylene glycol.
- Known unstable cardiopulmonary condition (e.g. acute myocardial infarction, acute coronary artery syndromes, worsening or unstable congestive heart failure, serious ventricular arrhythmias).

**Table S2. Primers and probes used for ddPCR assays**

| Variant | TERT C228T | TERT C250T | IDH1 R132H |
|---|---|---|---|
| **Mutant probe** | (6-FAM) 5'-AGCCCCTTCCGGGCCCTCCCA-3' (BHQ1) | (6-FAM) 5'-CGTCCCGACCCCTTCCGGGT-3' (BHQ1) | (6-FAM) 5'CATCATAGGTCATCATGCTTATGGGG -3' (BHQ1) |
| **Wild type probe** | (HEX) 5'-AGCCCCCTCCGGGCCCTCCCA-3' (BHQ1) | (HEX) 5'-CGTCCCGACCCCTCCCGGGT-3' (BHQ1) | (HEX) 5'CATCATAGGTCGTCATGCTTATGGGG-3' (BHQ1) |
| **Forward primer** | 5'-TCCAGCTCCGCCTCCTCC-3' | 5'-TTCCAGCTCCGCCTCCTCC-3' | 5'-GGCTTGTGAGTGGATGGG-3' |
| **Reverse primer** | 5'-AGCGCTGCCTGAAACTCGC-3' | 5'-AGCGCTGCCTGAAACTCGC-3' | 5'-ACACATACAAGTTGGAAATTTCTGG-3' |

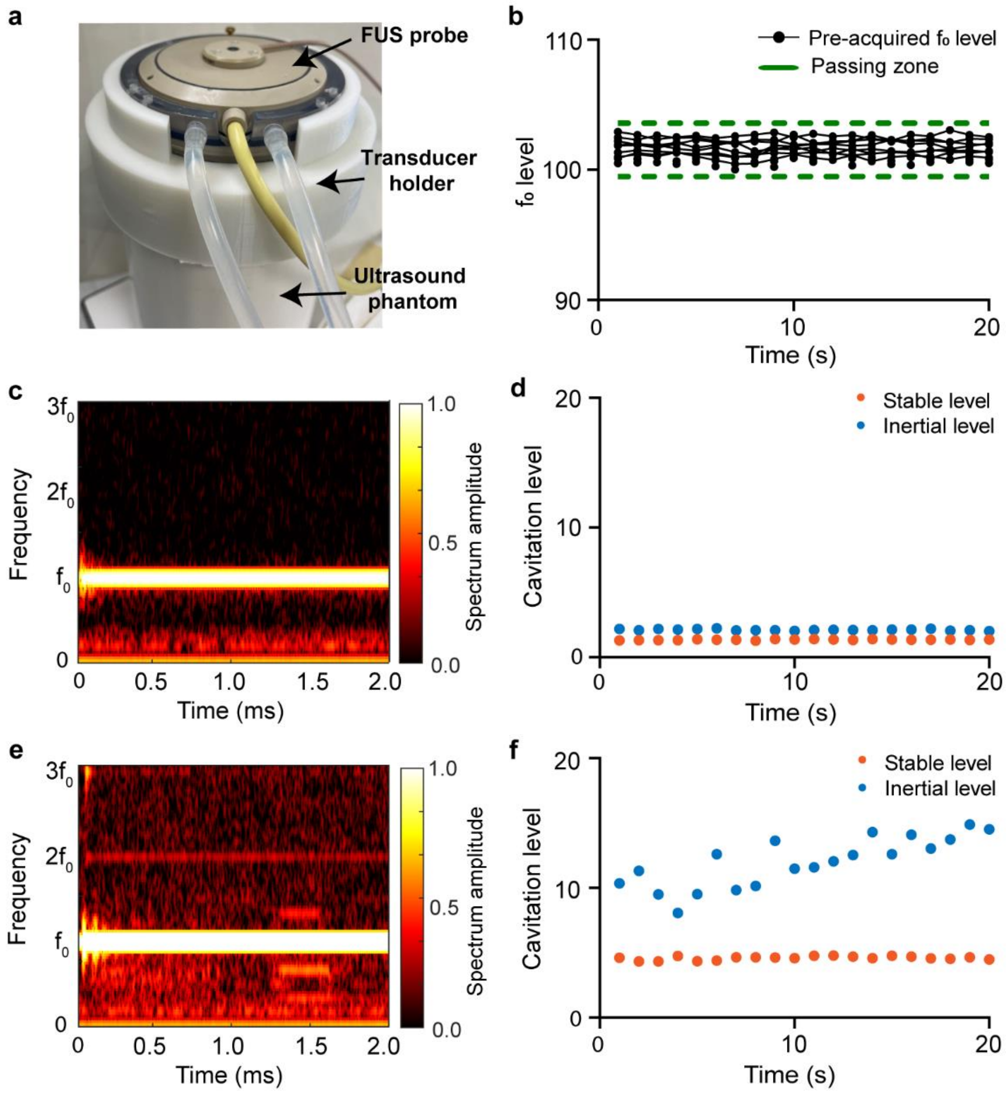


**Figure S1. Sonobiopsy device quality assurance tests. (a)** Setup for quality assurance check of the FUS transducer using a commercial ultrasound phantom (CIRS Technology, Norfolk, VA USA) with 3D printing transducer holder. FUS sonication was performed at 0.3 MPa (free-field acoustic pressure) on a commercial ultrasound phantom with a center frequency of 650 kHz, a pulse repetition frequency of 1 Hz, a pulse length of 10 ms (i.e., duty cycle: 1%) for 20 s. The acquired signals were analyzed in the frequency domain and the amplitude of center frequency (i.e., $f_0$ level) was compared with the pre-acquired $f_0$ levels to determine if the FUS probe has consistent output. **(b)** Pre-acquired $f_0$ levels and the passing zone calculated by the mean of the pre-acquired $f_0$ levels ± tolerance range. The tolerance range was defined by the three times standard deviation of pre-acquired $f_0$ levels. The FUS probe was considered as passing quality assurance if the $f_0$ level acquired before the sonobiopsy procedure was located within the passing zone. **(c–f)** Quality assurance for acoustic coupling during sonobiopsy procedure by performing FUS sonication before the injection of microbubbles. Although hairs were shaved before FUS sonication, a short length of hairs remained, which could trap air bubbles. Bubbles trapped in the FUS probe water bladder and ultrasound gel could reflect the ultrasound wave and decrease the delivered ultrasound energy in the brain. A representative example of good acoustic coupling shows mainly signal at $f_0$ **(c, d)**. A presentative example of bad coupling displays the presence of sub-harmonic signals and the increase of stable cavitation level **(e, f)**. The coupling procedure needs to be repeated to obtain good coupling before the sonobiopsy procedure.

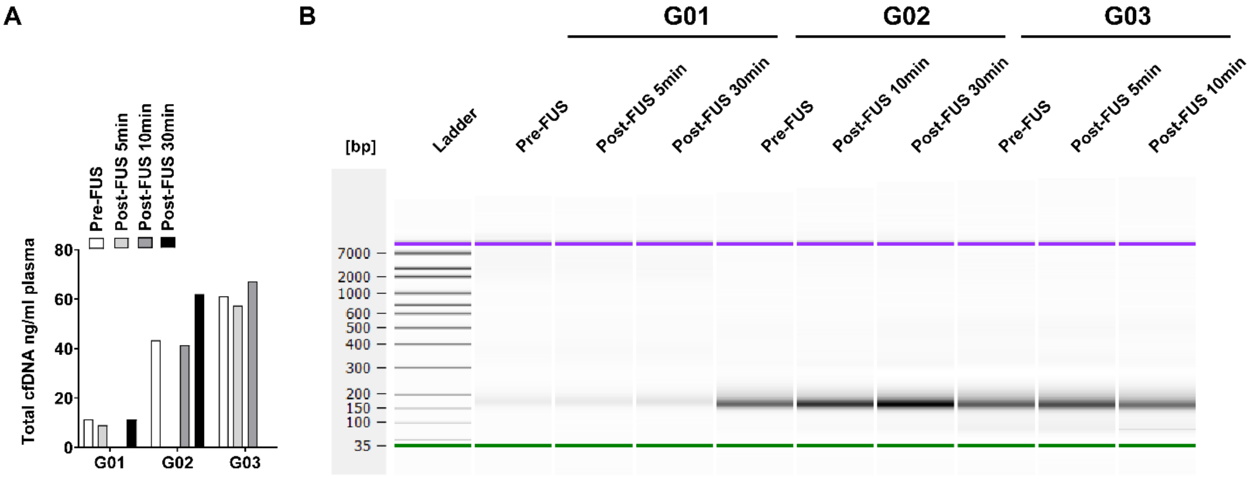


**Figure S2. Plasma cfDNA quantity analysis.** (A) Total plasma cfDNA concentration was elevated after sonobiopsy. (B) Bioanalyzer DNA gel analysis of plasma cfDNA used for personalized tumor-informed ctDNA assays and ddPCR assays. The peak size around 160bp representing mononucleosomal cfDNA clearly presented in each cfDNA sample.

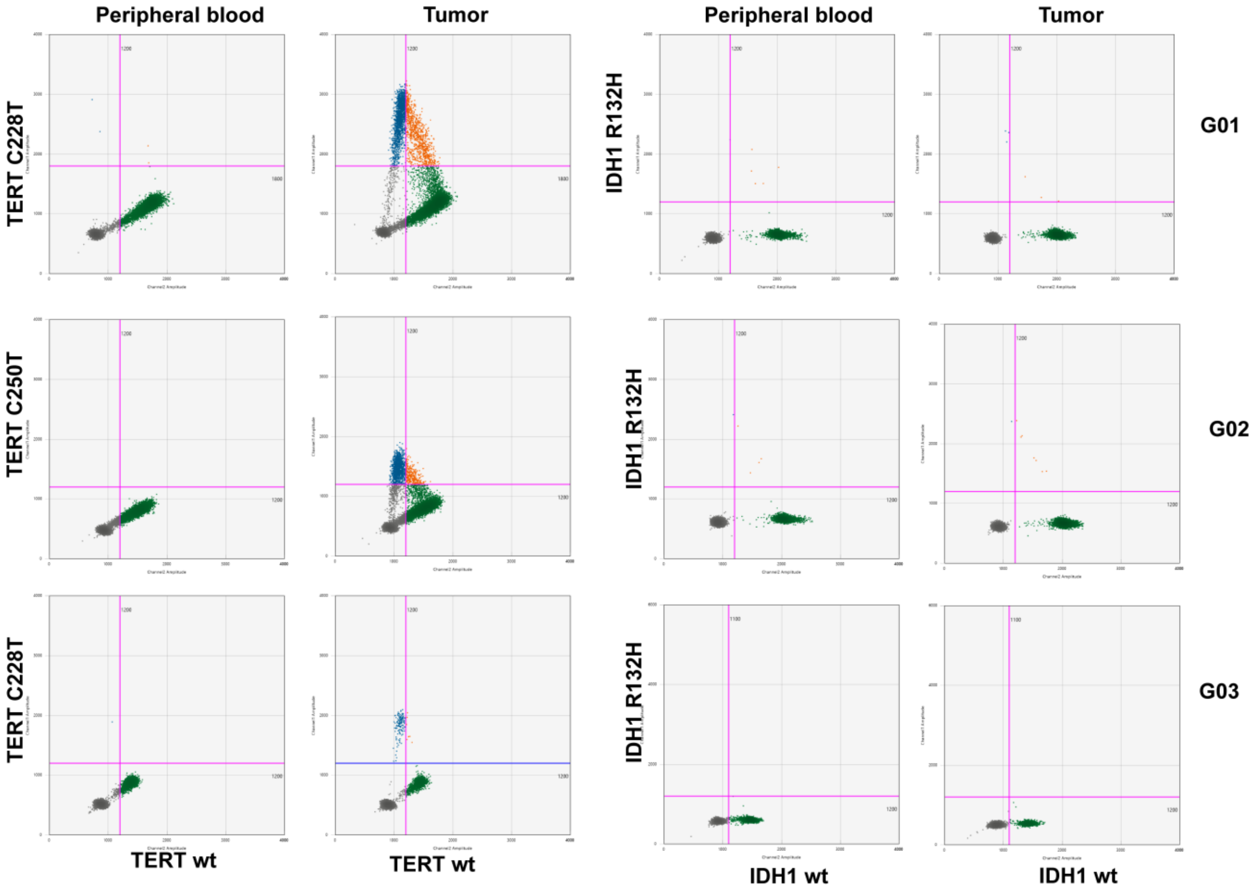


**Figure S3. Evaluation of TERT promoter and IDH1 mutation for three patients with ddPCR assays.** All three patients are positive for TERT promoter mutation and negative for IDH1 R132H mutaion.

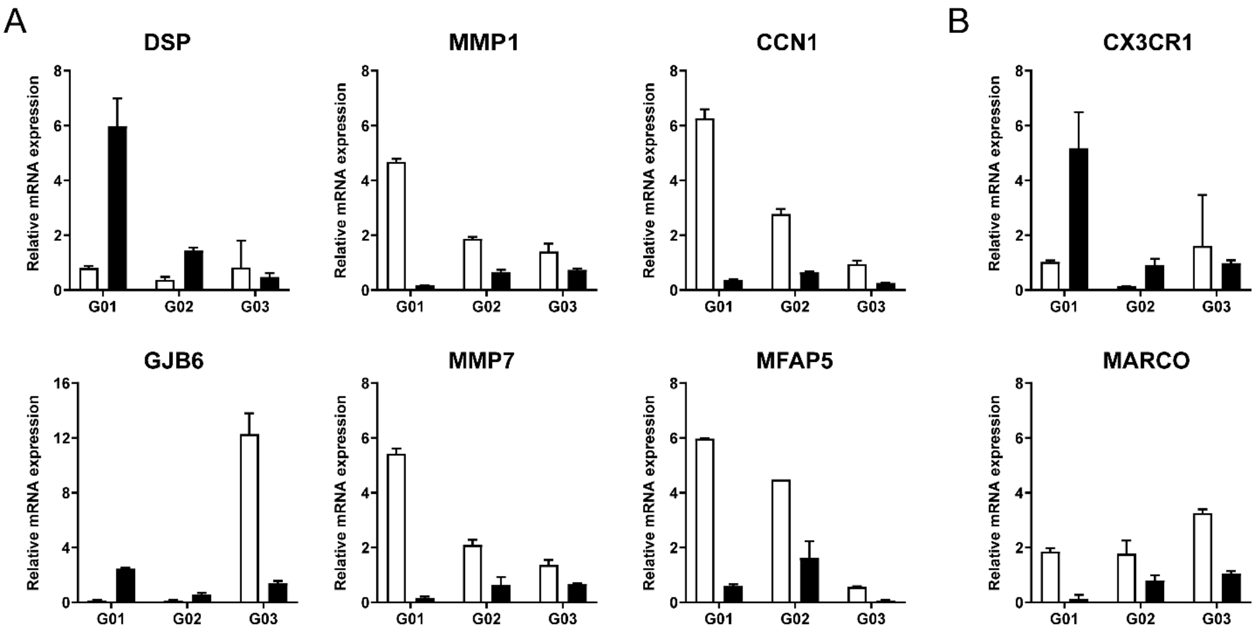


**Figure S4. Analysis of gene expression changes by FUS sonication using qRT-PCR.** (A) The relative mRNA expression for genes associated with cellular interactions after FUS sonication. The error bars show the standard deviation for the replicates in qPCR. (B) The relative mRNA expression for genes associated with inflammatory response after FUS sonication.
